# Revisiting a null hypothesis: exploring the parameters of oligometastasis treatment

**DOI:** 10.1101/2020.08.10.20172098

**Authors:** Jessica A. Scaborough, Martin C. Tom, Michael W. Kattan, Jacob G. Scott

## Abstract

In the treatment of patients with metastatic cancer, the current paradigm states that metastasis-directed therapy does not prolong life. This paradigm forms the basis of clinical trial null hypotheses, where trials are built to test the null hypothesis: patients garner no overall survival benefit from targeting metastatic lesions. However, with advancing imaging technology and increasingly precise techniques for targeting lesions, a much larger proportion of metastatic disease can be treated. As a result, the life-extending benefit of targeting metastatic disease is becoming increasingly clear. In this work, we suggest shifting this qualitative null hypothesis, and describe a mathematical model which can be used to frame a new, quantitative null. We begin with a very simple formulation of tumor growth, an exponential function, and illustrate how the same intervention (removing a given number of cells from the tumor) at different times effects survival. Additionally, we postulate where recent clinical trials fit into this parameter space and discuss the implications of clinical trial design in changing these quantitative parameters. Our model shows that while any amount of cell kill will extend survival, in many cases the extent is so small as to be unnoticeable in a clinical context or out-weighed by factors related to toxicity and treatment time. Recasting the null in these quantitative terms will allow trialists to design trials specifically to increase understanding of what circumstances (patient selection, disease burden, tumor growth kinetics) can lead to improved OS when targeting metastatic lesions, rather than whether or not targeting metastases extends survival for patients with (oligo-)metastatic disease.

## Introduction

In the treatment of patients with metastatic cancer, the current paradigm states that targeted treatment of metastatic lesions does not prolong life. This paradigm forms the basis of clinical trial null hypotheses, where trials are built to test the null hypothesis: patients garner no overall survival (OS) benefit from targeting metastatic lesions.

The development of distant metastases is the forerunner of cancer-related death (1–3). A Hallmark of Cancer, the dissemination of cancer cells from their origin to distant sites results from a complex cascade of biological events, which may subsequently allow for even more efficient tumor propagation (4–6). Eradicating the body of as much metastatic disease as feasibly possible to halt said process is a natural inclination. Yet, historically, a guiding principle in treating cancer has been that targeting metastatic lesions leads to poor outcomes, because the treatment is either too late or too morbid. However, with advancing imaging technology and increasingly precise techniques for targeting lesions, a much larger proportion of metastatic disease can be treated (7). As a result, the life-extending benefit of targeting metastatic disease is becoming increasingly clear.

Metastatic stage is typically described as a binary variable in a clinical setting, either present or not (M0 or M1), although certain cancer subtypes (e.g. colon, prostate) now have more gradiation in classifying a patient’s metastatic stage (8). The term “oligometastatic state” was first described in 1995 as an intermediary between localized and widespread metastatic disease where metastasis-directed treatment has the potential to be curative (9). Since then, results from several exploratory studies and randomized controlled trials using metastasis-directed therapy in such patients have accumulated to support its existence (10, 11).

Consensus definitions have since been proposed to further refine subgroups of oligometastasis (12–14). For example, the distinction between oligometastatic disease at presentation versus the development of oligometastatic disease following definitive treatment of non-metastatic cancer have been designated “synchronous oligometastases” and “metachronous oligorecurrence,” respectively. “Oligoprogression” describes growth of few metastases in the setting of otherwise stable (or responsive) disease whilst undergoing systemic therapy, and “oligopersistence” is characterized by having several lesions which have a poorer response to systemic therapy than others. Intra-patient heterogeneity often complicates diagnostics even further, where some lesions respond to therapeutics while others persist. These designations (and many more not listed) underscore the complexity with which researchers and clinicians are coming to understand this disease state.

In addition to refining the term “oligometastatic,” clinicians have examined the benefit of treating patients with oligometastases (28, 29). The implicit null hypothesis of these investigations, that targeting metastatic disease does not provide a life-extending benefit, stems from the current paradigm of metastatic cancer treatment. Table 1 summarizes the results of some of these recent phase II and III clinical trials, demonstrating that this null hypothesis is frequently (but not always) refuted. Even accounting for known positive publication bias (30, 31), there is substantial evidence that supports a changing paradigm in the treatment of oligometastatic patients. However, despite many studies showing a significant increase in overall survival (OS) when metastatic lesions are targeted, the null hypothesis in ongoing clinical trial planning has not changed.

**Table 1.** A summary of clinical trials that examine the benefit of providing local treatment to patients with oligometastases.

| Citation | CT Phase | Primary Location | Results | Description |
| --- | --- | --- | --- | --- |
| (15, 16) | II | NSCLC | Positive for PFS and OS | In the Gomez et al. trial, treating oligometastases ($\leq 3$ non-primary lesions) demonstrated significant improvement in PFS and OS compared to maintenance therapy alone. |
| (17) | II | NSCLC (EGFR/ALK negative) | Positive for PFS | In the trial by Iyengar et al., targeting the primary with radiotherapy and oligometastases with SBRT followed by maintenance chemotherapy provided significantly improved progression-free survival compared to maintenance chemotherapy alone. |
| (18, 19) | II | Variety | Positive for OS | In the “SABR-COMET” trial, treating all sites of oligometastatic cancer with SABR demonstrated significantly improved OS compared to standard palliative treatment. |
| (20) | II | Prostate (hormone sensitive) | Positive for composite of progression metrics | In the “ORIOLE” trial, treating all sites of oligometastases with SABR led to improved outcomes measured by 6-month rate of progression (by PSA, imaging, symptoms, androgen-deprivation therapy initiation, and survival) when comparing to observation alone. |
| (21) | II | Prostate | Positive for ADT-free survival | In the “STOMP” trial, in patients with metachronous oligometastasis, using metastasis-directed therapy (SBRT or surgery) provided longer ADT-free survival compared to surveillance alone. |
| (22, 23) | II | CRC | Positive for OS | In the “EORTC 40004” trial, treating liver metastases ($<10$, no extrahepatic disease) with RFA, systemic treatment, and +/- resection led to long-term OS improvement compared to systemic treatment alone. |
| (24) | II | ES-SCLC | Positive for PFS, negative for OS | In the “RTOG 0937” trial, treating oligometastases with PCI and consolidative radiotherapy to both the chest and metastases did not improve OS and did delay progression, compared to PCI alone. |
| (25) | III | Prostate | Positive for PSA progression, negative for OS | In the “HORRAD” trial, in patients with metastases to the bone (any amount), providing radiotherapy to the prostate along with ADT did not improve OS and did improve time to PSA progression, compared to ADT alone. Exploratory subgroup analysis suggested patients with $\leq 4$ bone metastases may benefit from prostate radiotherapy. |
| (26) | III | Prostate | Negative for OS in complete group, positive for OS in patients with lower metastatic burden | In Arm H of the “STAMPEDE” trial, radiotherapy to the prostate did not improve OS in unfiltered cohort of patients, compared to lifelong ADT. However, in a pre-specified subgroup analysis, significant OS improvement was observed among those with lower metastatic burden. |
| (27) | III | Nasopharynx | Positive for PFS and OS | In a trial by You et al., the addition of locoregional radiotherapy to the primary improved OS and PFS compared to chemotherapy alone in patients with (oligo- and poly-) metastatic nasopharyngeal carcinoma. |

In this work, we suggest shifting this qualitative null hypothesis, and describe a mathematical model which can be used to frame a new, quantitative null. We begin with a very simple formulation of tumor growth, an exponential function, and use it to show that while any amount of cell kill will extend survival, in many cases the extent is so small as to be unnoticeable in a clinical context or out-weighed by factors related to toxicity and treatment time. Recasting the null in these quantitative terms will allow trialists to design trials specifically to increase understanding of what circumstances (patient selection, disease burden, tumor growth kinetics) can lead to improved OS when targeting metastatic lesions, rather than determining whether targeting metastases can extend survival for patients with (oligo-)metastatic disease. We purposely begin with the most simplistic possible mathematical model, considering only total disease burden and doubling time. We do not consider complexities such as space, metastatic locations/connectedness (32), immune interactions or any heterogeneities – all things which could be considered in future iterations, but which make the model less generalizable. Finally, a sensitivity analysis is performed to confirm that our findings are consistent using alternative ordinary differential equations (ODEs) that may be used to model tumor growth (33).

Due to its breadth, the current qualitative null hypothesis may be incorrectly accepted or rejected without a quantitative model to help design optimal patient and treatment parameters. Numerous qualitative and quantitative prognostic factors exist to help identify patients with metastatic disease which is likely to follow a relatively indolent course. For example, with slower disease progression, patients are more likely to derive greater benefit from aggressively targeting their metastases. Other characteristics include the number of lesions and organs involved, the time course of presentation and progression, tumor histology, patient innate and adaptive immunity, and various biological features (34). It is crucial that we parse through which of these patient characteristics can meaningfully affect treatment outcomes in the setting of oligometastasis. By rethinking the null hypothesis of metastatic cancer treatment, research efforts can better serve our patients by bringing a deeper understanding of how well treatment works, who it works best for, and when it is most efficacious, rather than continually testing the implicit null hypothesis.

## Model

### Modeling Tumor Growth using an Exponential Function

Beginning with a very simple model of tumor growth, an exponential function, we will explore the effect of treatment in scenarios with different growth rates, treatment effectiveness, and timing of the intervention. While this overlooks many of the realities of real human cancers, such as spatial, intra- and inter-tumoral (35–37) heterogeneity, it captures many of the essential aspects of growth (38). Furthermore, in the absence of other specific knowledge, general arguments can be expounded upon, but additional undetermined complexities can severely limit generalizability. Let us then model a tumor of size (cell number), *N*, beginning with a single cell, and a growth rate, *r*, as follows:

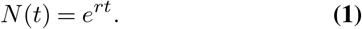

A growth curve built using Equation 1 is displayed in Figure 1 as the black line, denoted “Untreated.” The threshold tumor burden (an arbitrary number of *N* = 100 for illustrative purposes) which leads to patient death, *N*_*T*_, is represented by the horizontal black dashed line in Figure 1. Next, we will assume that a given intervention (e.g. stereotactic body radiation therapy (SBRT), or metastasectomy) is given at some time (e.g. upon detection of a metastasis). The total tumor burden at the time of this treatment is denoted as *N*_*d*_ and the number of cells killed is denoted as *N*_*c*_ cells; note, this requires 0 ≤ *N*_*c*_ ≤ *N*_*d*_.

**Fig. 1.**
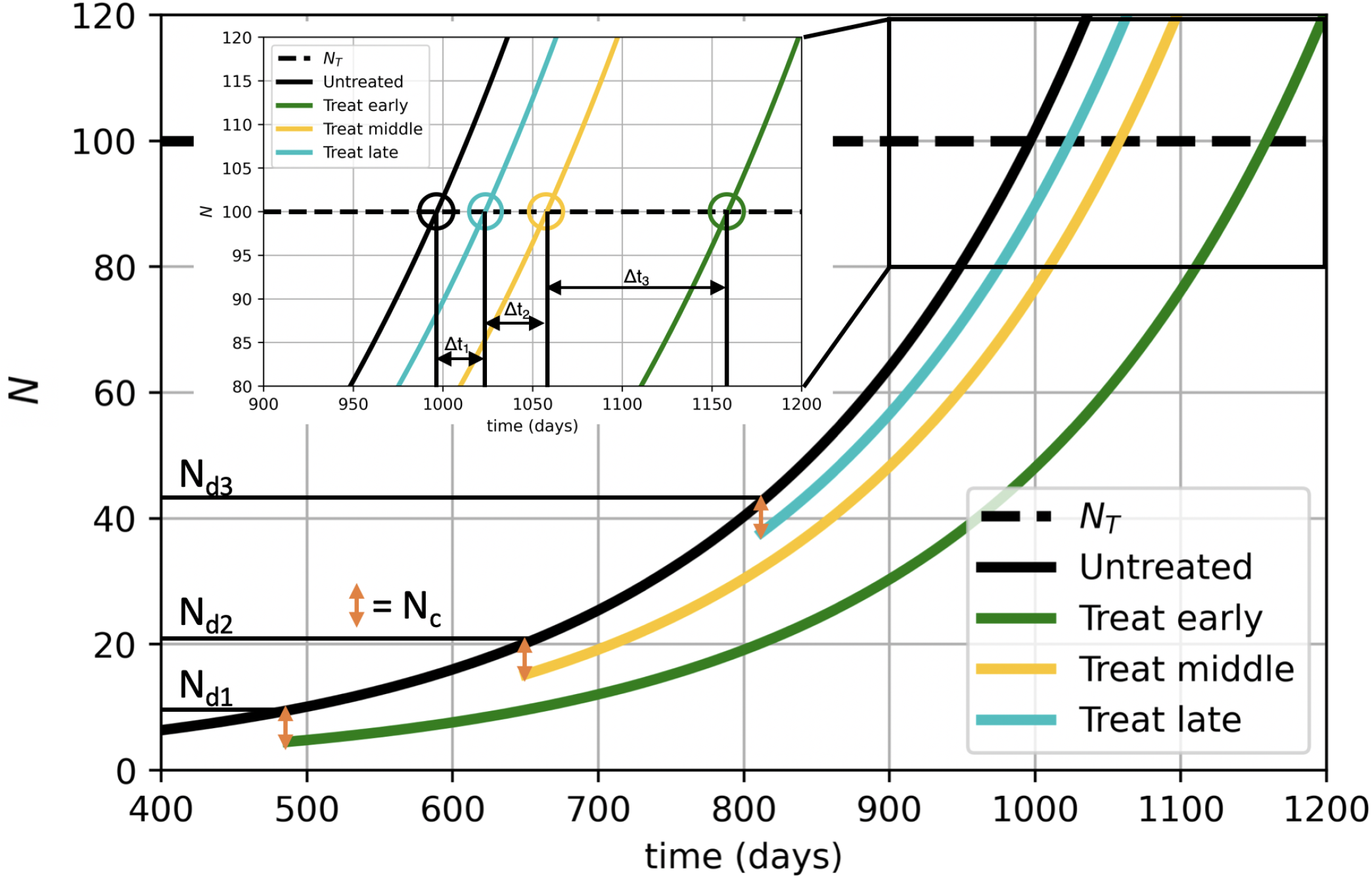
Change in OS is modulated by when an oligometastasis-directed intervention occurs and the effectiveness of the intervention. We plot an illustrative exponential growth curve from Equation 1 in black. At three different times, we subtract *N*_*c*_ cells from the curve to simulate an oligometastasis-directed intervention (orange markers), and the tumor continues to grow at the original rate from the new size. These subsequent tumors then grow and eventually intersect an arbitrary threshold cell (a surrogate for maximum tolerated disease burden) number (*N*_*T*_ - dashed horizontal line), there we can then determine the change in survival (vertical black lines, inset). The change in this time represents the Δ *t* for each intervention. n.b. These are not realistic parameters, but instead serve to illustrate the (qualitatively conserved) phenomenon.

To illustrate how the same intervention (removing *N*_*c*_ cells from the tumor) at different times effects our measure of survival, we plot several growth curves together in Figure 1. The time when each of these curves reaches *N*_*T*_ is the time of death (*t*_*d,x*_). The difference (Δ *t*) between the unperturbed time of death (*t*_*d*,1_) and each subsequent example intervention (e.g. Δ *t* = *t*_*d*,2_ − *t*_*d*,1_) is the increase in survival. We note that the earlier the intervention occurs (smaller *N*_*d*_), the greater the Δ *t* and, therefore, increase in survival. This is also true if we kill more cells (i.e. *N*_*c*_ increases).

While Figure 1 considers how a single intervention will effect the “same” tumor, Supplementary Figure 1 explores the effect of altering tumor growth rate, *r*, on Δ *t* after the same intervention. This figure adds a faster tumor growth curve, in addition to the curve seen in Figure 1. The same intervention (removal of *N*_*c*_ cells) occurs at the same time points as the slower curve, yet the faster growing tumor has a smaller resulting changes in survival time (Δ*tf*) compared to the slower growing tumor (Δ *ts*).

Next, we will examine the analytical relationship between the change in survival (Δ*t*) to the other parameters (*r, N*_*c*_, *N*_*d*_). This requires examining two tumor growth curves, one with unperturbed growth starting at *N*_*d*_ and the other with perturbed growth beginning at (*N*_*d*_ − *N*_*c*_). In other words, the perturbed curve will have the same growth characteristics as the unperturbed curve, but it will have *N*_*c*_ cells removed as “treatment.” Then, we will calculate the offset of time between the two curves when they reach *N*_*T*_, i.e. Δ *t*.

Graphically, we are asking how large the difference on the time axis is between where the treated and untreated curves intersect with *N*_*T*_ (the black dashed line), denoted by colored circles in Figure 1 and Supplementary Figure 1. Mathematically, we find the difference between *t*_*d*,1_ and *t*_*d*,2_: i.e. Δ *t* when 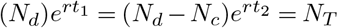. This relation is:

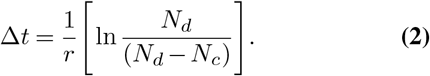

The observations from before are maintained: slower growing tumors (smaller *r*), more effective interventions (increasing *N*_*c*_), and lower burden at time of treatment (lower *N*_*d*_) make for a larger survival benefit, as we have intuited. Additionally, it is important to note that Δ *t* is not dependent on the threshold chosen for *N*_*T*_, where tumor burden leads to death.

Given the intuitive nature of these results, one may question what the value of such a model is. First, this model allows for the quantitative exploration of what was previously an exclusively qualitatively described phenomenon. This allows for formal interrogation of the individual values of each parameter, a crucial step in quantitative reasoning during clinical trial design. In doing so, a framework for parameter estimation can help trialists perform sensible power calculations. This would require measuring distributions of each of these parameters as it is clear that heterogeneity (and uncertainty) exists in each. Further, this would allow for error propagation in addition to power calculations. With recent work trying to incorporate toxicity into survival analyses in radiation oncology (39), we have the opportunity to formally probe the balance between benefit and harm in this setting. Most importantly however, it will remove the confusion created when we test a qualitative null that is likely neither able to be rejected or upheld given the sensitivity to the noise inherent in clinical data.

### Sensitivity Analysis with Alternative Tumor Growth Models

In order to assess the consistency of our findings, we examined the seven ODE models used in Murphy et al., 2016 (33), where they fit each model using 14 time points from a naked mouse xenograft experiment by Worschech et al., 2009 (40). The models included in this analysis are as follows: exponential, Figure 2A shows all seven models (including the exponential model) built using the parameters denoted by Murphy et al (33). Using these models, Figure 2B denotes the Δ *t*, or change in OS, between the untreated growth curves and growth curves with the same intervention at early, middle, and late timing. Here, we see that all models show that the same intervention has a greater benefit the earlier it is performed. Each model with untreated, early, middle, and late interventions is individually plotted in Supplementary Figure 2B-H, while Supplementary Figure 2A shows all untreated models together for comparison. Although not all models have as extreme of a difference as seen in the exponential model, this quantitative trend, which is the main thrust of this study, remains consistent.

**Fig. 2.**
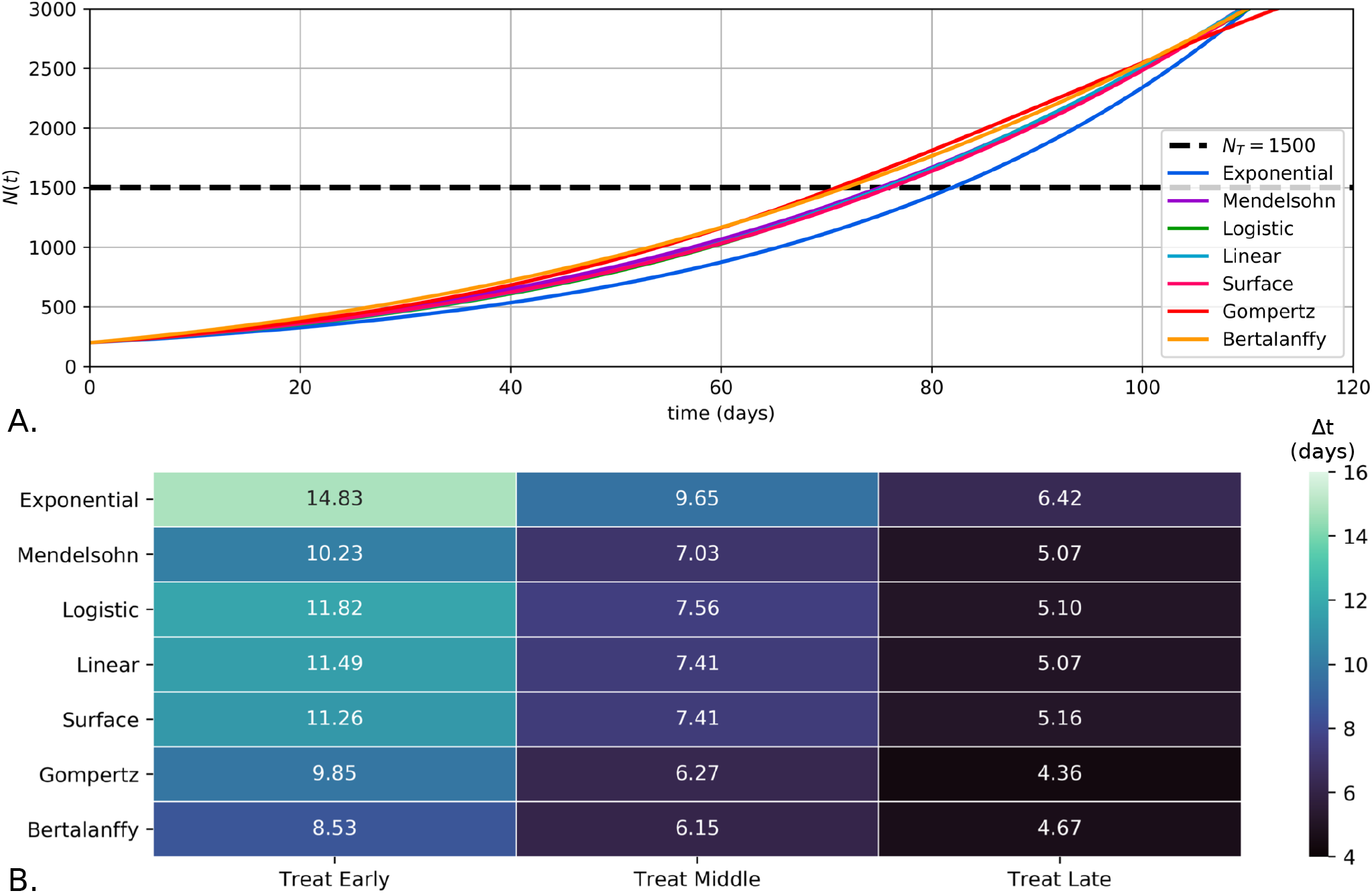
Across seven ODE tumor growth models, earlier intervention creates a larger improvement in OS. Models were produced using the parameters denoted by Murphy et al (33), where the seven models were fit to 14 timepoints of xenograft tumor growth data from Worschech et al (40). **A**. A comparison of the seven growth curves with no interventions built with various ODE models. Individual plots for each model and three intervention time points may be found in Supplementary Figure 2. **B**. A heatmap demonstrating the change in OS (Δ *t* days) for the same intervention (*N*_*C*_ = 100) at three different time points for each ODE model. Each heatmap entry is annotated with the exact change in OS for the given model and intervention timing. Treat early, treat middle, and treat late denote the intervention occurring at 20, 35, and 50 days, respectively.

### Parameter Sweep of Exponential Growth Model

Figure 3 demonstrates a benefit of using a quantitative model with a sensitivity analysis to help us better understand the areas of the (very simplified) parameter space, a range of possible parameter values, where the greatest opportunities lie. Given that this is a simple exponential relation, the change in survival is monotone (always up or down) in each parameter. However, as the tumor growth curves are non-linear, we chose to plot the sensitivity analysis on a log-log plane to improve the visualization of changes in parameter values.

**Fig. 3.**
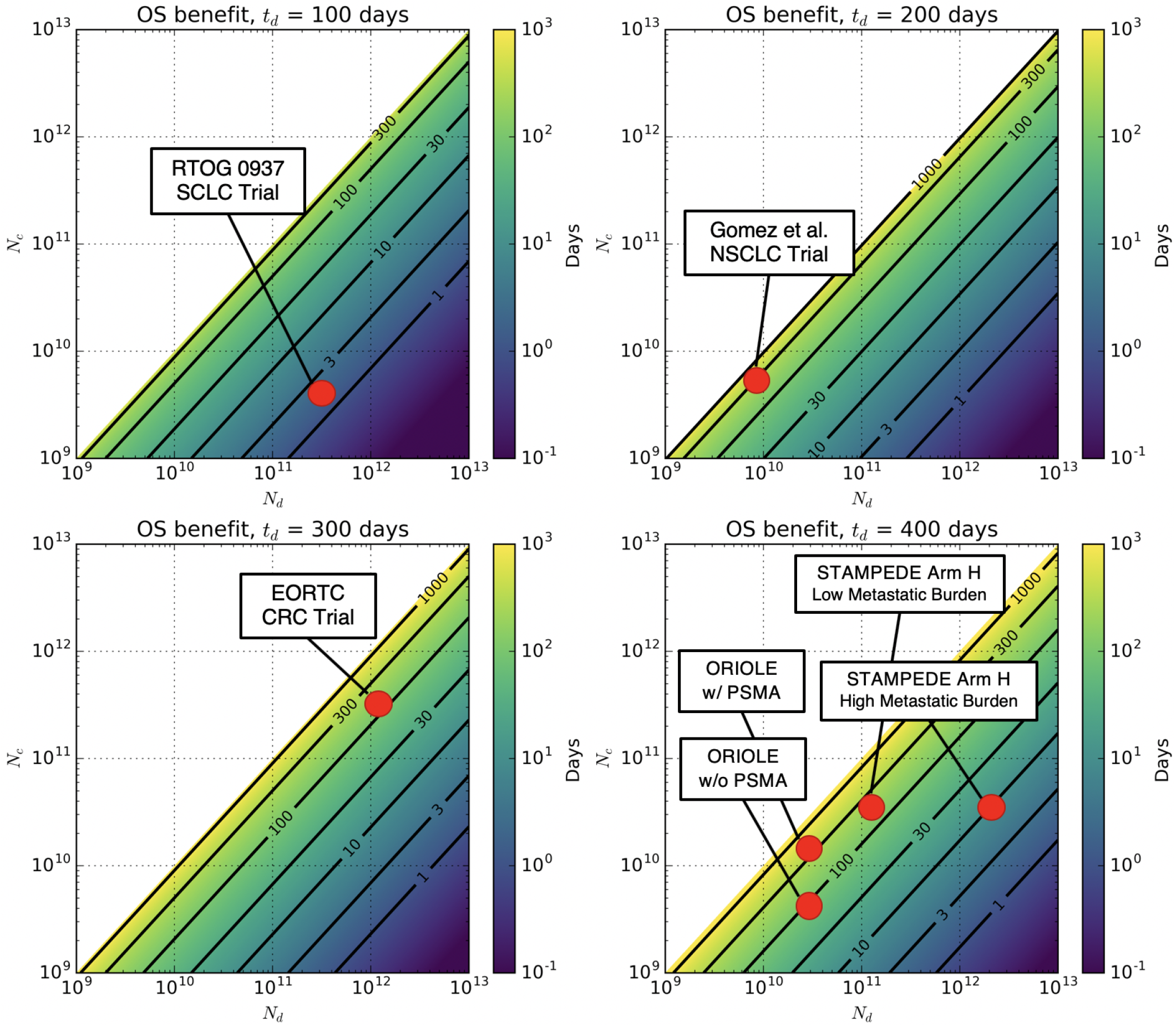
The benefit of oligometastasis-directed therapy depends monotonically on the amount of cells killed, the tumor burden, and tumor doubling time. We plot four orders of magnitude of both *N*_*c*_ and *N*_*d*_ on a log scale. The color represents the predicted number of days of OS (PFS in the ORIOLE trial) benefit for each combination of *N*_*c*_ and *N*_*d*_. Each of the four subplots represents a different “intrinsic” biology, modeled by different tumor doubling times. A *t*_*d*_ of 100, 200, 300, and 400 days corresponds to a growth rate, *r*, of 0.0069, 0.0035, 0.0023, and 0.0017, respectively. Contour lines are shown for ease of comparison. A selection of trials from Table 1 are represented by red circles based on estimations of *N*_*d*_, *N*_*c*_, *r*, and *t*_*d*_ for each trial.

As we do not currently have known values for these parameters, exploring a large sweep of values can be instructive. We consider a continuous range for *N*_*c*_ in [0, *N*_*d*_ where *N*_*c*_ = 0 represents no intervention and *N*_*c*_ = *N*_*d*_ represents a cure. In these cases, Δ *t* = 0 and Δ *t* = ∞, respectively. In Figure 3, we will consider four discrete examples of values for *r*, as this parameter’s effect is monotone (where a case with lower *r* always derives more benefit from oligometastasis-directed therapy than a case with higher *r*). It is also important to note that this parameter is likely modifiable with chemo- or targeted-therapy: something we do not consider here, but would be a straightforward extension. This example will consider growth rates which correspond to tumor doubling times of 100, 200, 300 and 400 days. These could represent tumors such as small cell lung cancer in the fast extreme or prostate cancer in the slow extreme. Figure 3 shows this analysis, with isoclines shown in black to denote lines of equal effect. These curves demonstrate that any increase in *N*_*c*_ (more cell kill per intervention, “up” on the y-axis) and/or decrease in *N*_*d*_ (earlier intervention, “down” on the x-axis) increases the OS benefit. It is interesting to note that the movements (i.e. *N*_*c*_ up and *N*_*d*_ down) mirror the historical trend: improvements in detection of oligometastasis via anatomic or functional imaging have slowly pushed *N*_*d*_ lower over the years and the ability to safely (using SBRT or minimally invasive surgery with continually lowering toxicity) target larger and larger lesions (increasing *N*_*c*_) has increased. This “creep” of these values is one reason why the need for a recasting of the null hypothesis is becoming clear, and why the null was historically of greater clinical utility.

### Sample Size Calculation Informed by Exponential Growth Model

Finally, we performed theoretical sample size calculations (using the Sample Size Estimator from Wang et al., 2020 (41)) in four clinical scenarios: a fast growing tumor type which is detected early, a fast growing tumor type which is detected late, a slow growing tumor type which is detected early, and a slow growing tumor type which is growing late. Using Equation 2, we assume parameters which are congruent with each clinical scenario and calculate the change in survival time between the treated and untreated growth curves. These Δ *t*’s are then converted into hazard ratios (HR) relative to each other, with the largest change in survival time relating to the most extreme HR (0.50) and the smallest change in survival time leading to the HR closest to 1 (0.80). In Table 2, we see that as the HR approaches 1, the predicted sample size requirements become unreasonable for any oncology treatment clinical trial. This HR may occur in a trial with a fast growing tumor type (e.g. SCLC) that has widespread metastatic disease.

**Table 2.** Theoretical sample size calculations demonstrate that as the change in OS decreases, the hazard ratio (HR) gets closer to 1, and a larger sample size is predicted. Using Equation 2, _Δ *t*_, representing change in OS, was calculated for four clinical scenarios. This predicted change in OS is translated to an estimated hazard ratio to perform a sample size calculation. The sample size calculations assumed a type I error (*α*) of 0.05, power (1 − *β*) of 0.80, equal size of treatment arms, and a superiority margin of 0.20. Sample size was estimated using a resource by Wang et al., 2020 (41) (http://riskcalc.org:3838/samplesize/), which references Schoenfeld et al., 1981 (42) and 1983 (43) for the calculation of sample size in parallel randomized control trials for assessing superiority of a treatment using time-to-event.

| Tumor Parameters | Doubling Time (days) | Growth Rate ($r$) | $N_d$ | $N_c$ | $\Delta t$ (days) | HR | Sample Size |
| --- | --- | --- | --- | --- | --- | --- | --- |
| Fast growing tumor, early detection | 100 | 0.0069 | $10E10$ | $5E10$ | 100.0 | 0.60 | 82 |
| Fast growing tumor, late detection | 100 | 0.0069 | $10E11$ | $5E10$ | 7.4 | 0.80 | 14698 |
| Slow growing tumor, early detection | 400 | 0.0017 | $10E10$ | $5E10$ | 400.0 | 0.50 | 34 |
| Slow growing tumor, late detection | 400 | 0.0017 | $10E11$ | $5E10$ | 29.6 | 0.70 | 322 |

As seen in McClatchy III et al., 2020 (44) and XXXX et al., 2020 (39), a more technical method of assessing these theoretical sample size calculations would be to perform an *in silico* clinical triald. One would assume distributions for each parameter of interest (e.g. *r, N*_*d*_), sample each “patient” in the trial from these distributions, assess survival time for treated and untreated groups, and calculate the HR between the two groups. Even with this theoretical approach, however, additional measurements would be required to appropriately estimate the distributions of the model parameters. Post-trial data publication could provide a wealth of information to estimate these parameters. For example, tumor imaging both before and after treatment could assess *N*_*d*_ and *N*_*c*_, while serial tumor biopsies could be used to inform a distribution for tumor growth rate. Without these data, however, we believe that the simpler method of converting Δ *t* to HR shown in Table 2, provides an easier-to-interpret example of sample size calculations.

### Clinical correlation

In order to demonstrate how clinical trial design can explore the parameter space of this tumor growth model, we will review some recent clinical trials, which are also listed in Table 1. This discussion reviews illustrative examples, and is not an exhaustive list of all clinical trials which test the benefit of targeting oligometastases. For many trials, we will estimate where design falls in the parameter space of Figure 3, and discuss how trial design can test the effects of altering one or more parameters (i.e. *N*_*d*_ *N*_*c*_, or *r*).

In a phase II trial by Gomez et al., 49 patients with oligometastatic (≤ 3 metastases) non-small cell lung cancer (NSCLC) without progression after first-line systemic therapy were randomized to either maintenance systemic therapy/surveillance or local consolidative therapy (LCT) to all sites of residual disease via surgery or radiotherapy. After interim analysis demonstrated a substantial PFS benefit with LCT, the trial was closed early and allowed for crossover to the LCT arm (15). With additional follow up, and despite crossover, LCT was associated with improved OS of 41.2 months vs 17.0 months (16). We placed this trial in the top right subplot of Figure 3, due to the relatively fast growth rate of NSCLC, minimal tumor burden (≤ 3 metastases), and large *N*_*c*_ using radiotherapy or surgery.

The SABR-COMET study was a screening phase II trial which randomized 99 patients with oligometastatic disease (≤ 5 metastases) of various histologies with a controlled primary site to standard palliative therapy with or without stereotactic ablative radiotherapy (SABR) to all metastatic lesions. The primary endpoint was OS, which was initially improved with the addition of SABR from 28 months to 48 months (19). With additional follow up, results were even more substantial with a median OS of 50 months using SABR versus 28 months in the control arm (45). As this trial includes tumors of many histologies, we cannot place the positive results in a single subplot of Figure 3, but doing so *post-hoc* patient by patient would be illustrative. The SABR-COMET trial also utilized stratified randomization to ensure that strata of patients with 1-3 metastases and 4-5 metastases were balanced in treatment assignments. Stratified randomization helps balance treatment arm assignments between patient populations with known prognostic factors, and can reduce the risk of type I and II errors in trials with smaller sample sizes (<400 patients) (46). In relation to our model, by creating strata of the number of metastases, the SABR-COMET trial balanced based on tumor burden at the time of treatment (*N*_*d*_). In future, other trials may consider stratifying based on tumor growth rate (either inferred by tumor type or measured from serial tumor biopsies), the sensitivity of imaging techniques (*N*_*d*_ or *N*_*c*_), or the efficacy of two treatment types (*N*_*c*_).

In the phase II EORTC 40004 trial, 119 patients with fewer than 10 unresectable colorectal liver metastases and no extrahepatic disease were randomized to systemic therapy with or without local therapy using RFA (with or without resection). Although the primary endpoint of 30 month OS was not met, longer follow up led to improved OS with RFA from 40.5 months to 45.6 months (23). With a relatively slow growing tumor sub-type, a large *N*_*d*_, and a moderate OS benefit, we estimated this clinical trial to fall in the bottom left subplot of the model’s parameter space found in Figure 3.

The largest study was Arm H of the STAMPEDE trial, which was a phase III trial of 2061 patients with metastatic prostate cancer randomized to androgen deprivation therapy with or without definitive radiotherapy to the prostate. Prespecified subgroup analysis demonstrated no benefit to the addition of prostate radiotherapy among those with a high metastatic burden, defined as either visceral metastases or ≥ 4 bone metastases with ≥ 1 outside of the vertebral bodies or pelvis. However, in the group of 819 patients with a low metastatic burden, radiotherapy to the prostate improved three-year OS from 73 percent to 81 %. (26) In relation to our model, this is equivalent to assuming that the two groups (high and low metastatic burden) have different *N*_*d*_ at the time of treatment, but experience the same *N*_*c*_. It should be noted that unlike other trials discussed, local therapy was delivered only to the primary site, but not the metastatic sites, suggesting a benefit to cytoreduction. The estimated parameter space position of these two subgroups (high metastatic burden and low metastatic burden) is found in the bottom right subplot of Figure 3.

In the ORIOLE trial, patients with metachronous oligometastatic prostate cancer with ≤ 3 sites as detected by conventional imaging were randomized to surveillance or SABR to all sites (20). The primary endpoint was a composite of disease progression metrics at 6 months, which was improved with SABR at 19% versus 61% in the control arm. Interestingly, a subgroup of patients underwent advanced imaging with PSMA PET, which has demonstrated greater sensitivity in detecting prostate cancer metastases (putatively lowering *N*_*d*_) (47). Among those patients where all PSMA PET avid sites were treated, the 6 month progression rate was just 5% compared to 38% in those with untreated sites. This subgroup analysis further supports that advanced imaging can better identify metastases and treating all sites improves outcomes. By utilizing a more sensitive technology in detecting (and therefore targeting) metastases, we see that a greater *N*_*c*_ increases PFS, even if *N*_*d*_ remains the same. We estimate the parameter space for this subgroup analysis in the bottom right subplot of Figure 3.

Not all trials have demonstrated benefit to the addition of metastasis-directed therapy. For example, RTOG 0937 was a phase II study of 86 patients with extensive stage small cell lung cancer with at least a partial response to chemotherapy and 1-4 extracranial metastases who were randomized to prophylactic cranial irradiation with or without consolidative radiotherapy to the chest and all metastatic sites. The primary endpoint of one-year OS was not significantly different; 60% in the control arm and 51% in the consolidative radiotherapy arm (24). This negative result is estimated to be in the top left subplot of Figure 3, due to the rapid growth of SCLC. Here, this model could have still been useful by informing the sample size calculations (Table 2), given the parameter estimates of the trial’s patient population.

## Conclusions

In this work, we have used a simple exponential model of tumor growth to demonstrate why recent improvements in metastasis detection and treatment may allow us to reconsider the null hypothesis when treating patients with oligometastases. Specifically, more sensitive techniques to localize metastases (as seen with PSMA imaging) increases how many tumor cells are removed, *N*_*c*_, when considering patients at similar stages. When used for surveillance, these imaging techniques can decrease the tumor burden at the time of treatment, *N*_*d*_, while still increasing the efficacy of therapy, *N*_*c*_, potentially leading to clinically significant improved OS. Next, advancements in the ability to administer local therapy to all sites of disease with surgical resection, radiotherapy, and/or ablative procedures such as radiofrequency ablation (RFA) has allowed for more effective, precise eradication of metastatic lesions with reduced associated morbidity. Furthermore, novel immuno- and targeted-therapies can likely decrease the growth rate, *r*, of tumors.

A mathematical model provides the distinct advantage of testing quantitative hypotheses to optimize the treatment of patients with oligometastases. Parameter selection regarding number of oligometastases, measurements of tumor burden, and efficacy of treatment options can be examined with robust hypotheses born from simulated results. Additionally, with increased translation between the bench and bedside, some model parameters (e.g. *r*, tumor growth rate) may be inferred using serial tumor biopsies, *in vitro*, or *in silico* modeling. A deeper understanding of how these parameters affect outcome can improve trial design by allowing for rational prognostic strata criteria in stratified randomization or informing prior probabilities in a Bayesian clinical trial (46, 48, 49). Furthermore, Bayesian trial interim analyses can be enhanced with additional simulations using updated parameters as patient characteristics are observed or assessed over time. With a better understanding of the prognostic factors of the population enrolled in a trial, these interim analyses may be used to update prior probabilities, predict probability of success, and assess sample size requirements (48, 49).

It is important to note that the model demonstrated in this work is not a perfect representation of tumor growth and treatment, as it fails to consider intratumoral heterogeneity, metastasis location, and the inherent risks of treatment. However, because of its simplicity, this model provides a foundation exploring the current parameter space, while allowing researchers to add complexity as they see fit.

There are minimal published clinical trial results which support upholding the current paradigm in the treatment of oligometastases; however, this is likely due in part to publication bias where positive results are more likely to be published, not simply because this null hypothesis has always been rejected (30, 31). The clinical trials examined in this manuscript have necessarily sought to examine the fundamental idea that oligometastatic lesions should only be targeted for palliative care. Refuting this standard was crucial, as the earlier state of cancer imaging and treatment established that targeting oligometastases either occurred too late or caused too much harm. Yet, as quantitative models of tumor growth and the knowledge of how metastatic detection and treatment have evolved, we believe that clinical trials can now provide an even greater benefit by reconsidering the default null hypothesis and utilizing the quantitative principles of this mathematical model in trial design.

## Data Availability

All data and code used in this manuscript may be found at in the link provided.

https://github.com/jessicascarborough/oligo-null

## Code Availability

All code used to create mathematical models and figures in this manuscript may be found on GitHub at https://github.com/jessicascarborough/oligomet-null-hypothesis.

## ACKNOWLEDGEMENTS

JGS thanks his patients for providing him with motivation to push the boundaries of what we know. He would also like to thank the NIH for their support through NIH R37CA244613 and their generous Loan Repayment Program and the American Cancer Society for the Research Scholar Grant (Award number: 132691-RSG-20096-01-CSM). JAS was supported in part by NIH grant T32 GM007250.

## Supplementary Information

**Supplementary Fig 1.**
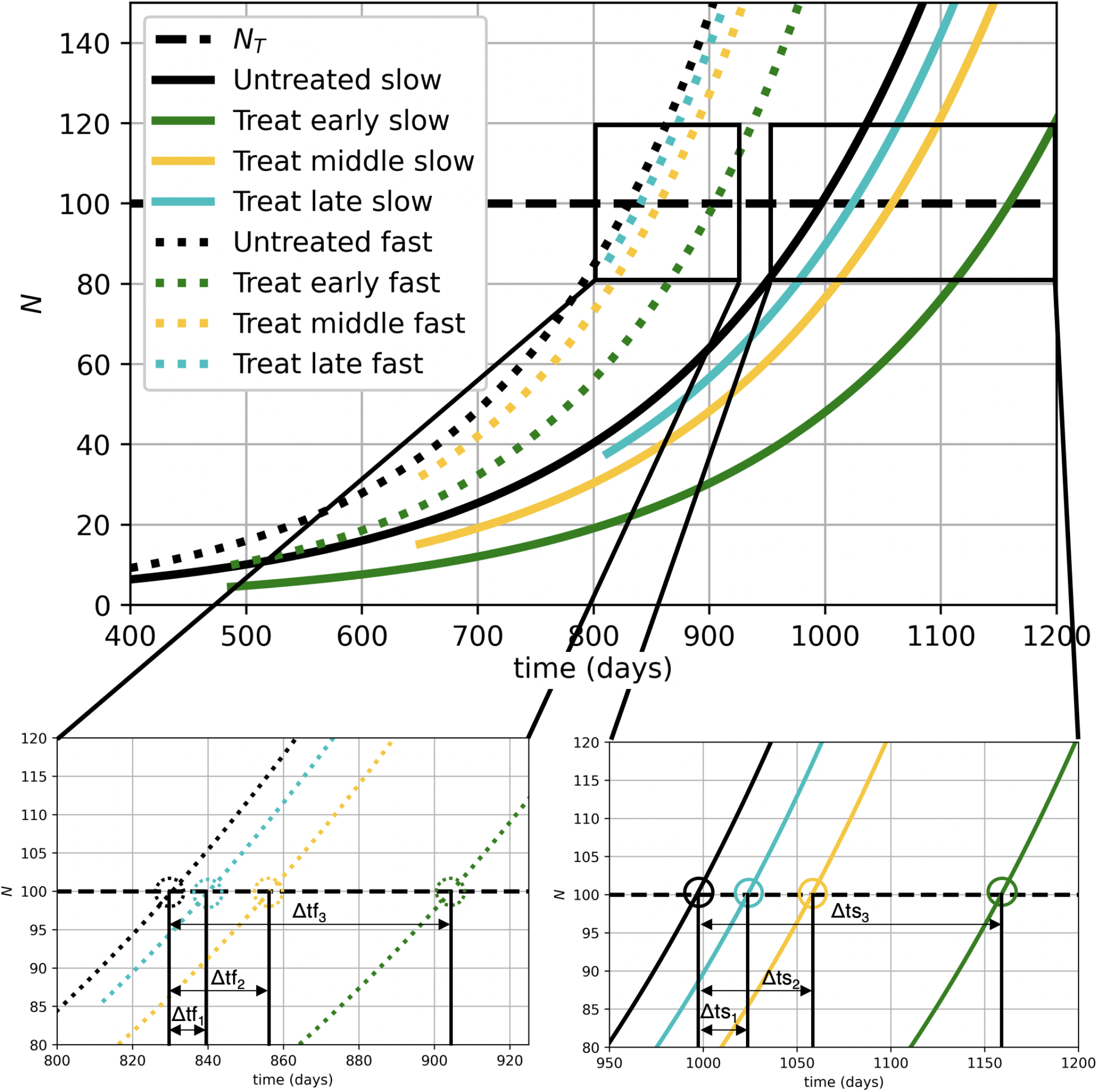
Change in OS is modulated by tumor growth rate, intervention timing, and intervention efficacy. **Top:** We plot two illustrative exponential growth curves from Equation 1 in black, using a faster (dotted line) and slower (solid line) growth rate, *r*. The slower growth rate is the same curves shown in Figure 1. At three different time points, we subtract *N*_*c*_ cells from the two curves to simulate an oligometastasis-directed intervention, and the tumor continues to grow at the original rate from the new size. These subsequent tumors then grow and eventually intersect an arbitrary threshold cell (a surrogate for maximum tolerated disease burden) number (*N*_*T*_ - dashed horizontal line). **Bottom:** We plot two expanded windows of the above plot, showing greater detail of the faster (left, dotted) and slower (right, solid) growth curves as they reach *N*_*t*_. In these plots, we can then determine the change in survival (vertical black lines). The change in this time represents the Δ *tf* and Δ *ts* for each intervention in the fast and slow curves, respectively. Notably, the x-axis for the faster (left, dotted) growth curves accounts for fewer days, despite having the same relative length as the x-axis for the slower (right, solid) growth curves. This was necessary in order to annotate the smaller Δ *tf* for the faster growth curves.

**Supplementary Fig 2.**
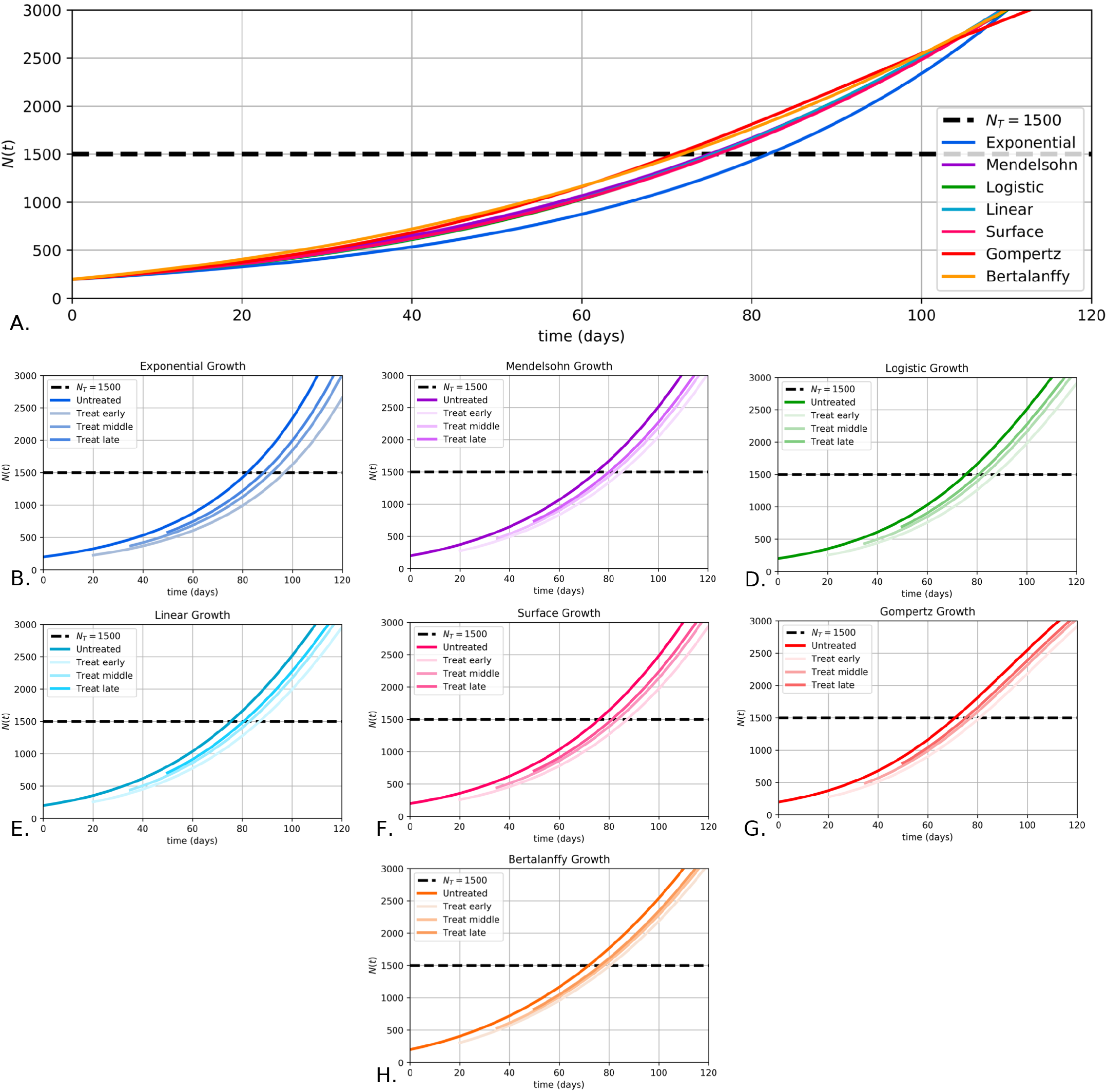
A visual representation across seven ODE tumor growth models, demonstrating that earlier intervention creates a larger improvement in OS. Models were produced using the parameters denoted by Murphy et al (33), where the seven models were fit to 14 timepoints of xenograft tumor growth data from Worschech et al (40). **A**. A comparison of the seven growth curves with no interventions built with various ODE models. **B-H**. Individual plots visually demonstrating the change in OS for three intervention times for all models included in A. The calculation of change in OS, is demonstrated in Figure 1 with the calculation of Δ *t*.

